# Defining suspected child maltreatment using International Classification of Diseases codes: A scoping literature review

**DOI:** 10.1101/2022.06.12.22276294

**Authors:** Yuerong Liu, Lindsay Terrell, Brianna Joyce, Julia Black, Samantha J. Kaplan, Elizabeth J. Gifford

**Affiliations:** Center for Child and Family Policy, Duke University, Durham, North Carolina; Sanford School of Public Policy, Duke University, Durham, North Carolina; School of Medicine, Duke University, Durham, North Carolina; Department of Pediatrics, Duke Children’s Primary Care, Duke University Medical Center, Durham, North Carolina; Margolis Center for Health Policy, Duke University, Durham, North Carolina; Children’s Health and Discovery Initiative, Duke University, Durham, North Carolina

**Keywords:** Child maltreatment, International Classification of Diseases (ICD), review

## Abstract

**Background:** Administrative medical and claims records are increasingly used to study prevalence of and outcomes for children exposed to child maltreatment. However, suspicion of child maltreatment is often under-documented in medical records using International Classification of Diseases (ICD) codes. Although researchers have developed strategies to more broadly capture the injuries, illnesses, and circumstances that are suggestive of maltreatment, there is no consensus on which codes to use for this purpose.

**Objective:** To systematically examine the types of research being conducted with ICD codes related to suspected maltreatment, summarize the methods used to identify the codes, and propose future direction.

**Methods:** We searched five electronic databases for studies that reported ICD codes suggestive of child maltreatment in any medical setting, included participants aged 0-18 years, and published in a peer-reviewed journal in English. Two reviewers independently screened the titles, abstracts, and the full texts. Data were synthesized in a qualitative manner.

**Results:** Thirty-seven studies met inclusion criteria. Most studies focused on determining the incidence or trends of maltreatment-related injuries or illnesses. Studies varied greatly in the codes used to identify suspected maltreatment. Only four articles reported on the validation of selected codes. ICD codes for transport accidents were the most frequently excluded co-occurring codes. Studies frequently relied on two seminal studies and two national-level guidelines.

**Conclusions:** Substantial heterogeneity existed in the ICD codes and methods used to identify suspected maltreatment. The age range for codes, excluded co-occurring codes, and validation of codes are areas future research should address. This review may reduce costs for future researchers so that they could choose optimal measures of suspected maltreatment from our summarized list of codes without reinventing the wheel. Our review provides a basis for the development of recommended guidelines in establishing uniform codes for suspected maltreatment that could promote public health surveillance and allow for more efficient and uniform policy or program evaluation.

## Introduction

Child abuse and neglect (maltreatment) is a serious public health problem with enduring deleterious effects on children (Jonson-Reid et al., 2012). Health care providers are instrumental in identifying and treating children -- especially young children -- who have experienced maltreatment (Jackson et al., 2015). As such, electronic medical records and health care claims data are key resources for cost effective maltreatment research at the population level. However, documentation of maltreatment in medical records is notoriously inconsistent across providers and often under-reported (Scott et al., 2009). Thus, researchers using administrative health care data have developed strategies to more broadly capture the experiences of children who were potentially exposed to maltreatment. Because no standardized guidelines exist to construct these broader definitions, this study aimed to summarize the various approaches used to identify and apply the International Classification of Diseases (ICD) codes suggestive of child maltreatment. By understanding how these ICD codes were used in a research context and documenting the various approaches which have been used to operationalize a latent construct, this study may reduce costs to future researchers who are trying to determine how to best measure suspected maltreatment.

The ICD system is an international standard for classifying causes of death and morbidity, as well as documenting and aggregating clinical diagnoses. The classification system includes specific diagnostic codes or external cause of injury codes for the documentation of child maltreatment and serves as a foundation for tracking of health trends globally (World Health Organization, 2021). Worldwide, child maltreatment is regularly under-documented in electronic health records (González-Izquierdo et al., 2010; McKenzie & Scott, 2011; Scott et al., 2009; Wu et al., 2015) owing to factors such as missed or unrecognized sentinel injuries or early signs of maltreatment (Sheets et al., 2013), lack of confidence or reluctance of medical professionals to report suspicious maltreatment to CPS (Flaherty et al., 2008; Schnitzer et al., 2011), and insufficient ICD codes for neglect-related conditions (Schnitzer et al., 2011). An estimated 11%-33% of children diagnosed with child maltreatment have been previously evaluated by a medical professional for signs and symptoms caused by maltreatment, but the diagnosis of maltreatment was missed (King et al., 2006; Letson et al., 2016; Oral et al., 2003; Quiroz et al., 2020; Thorpe et al., 2014). Undiagnosed or delayed diagnostics of child maltreatment may lead to chronic abuse and increased morbidity-mortality (King et al., 2006).

The ICD-10 nosology provides availability to document suspected maltreatment by distinguishing confirmed (e.g., ICD-10-CM code T74) and suspected (e.g., ICD-10-CM code T76) diagnosis of maltreatment (Feng et al., 2011), yet studies have indicated that the codes are still under-used (Hunter et al., 2021) and thus inadequately captured the breadth of suspected maltreatment. One study using retrospective chart review from a pediatric trauma center found that only 17% of suspected physical abuse cases identified by chart review were captured using ICD-10 diagnostic codes for suspected abuse (Durand et al., 2019). Despite this, the injuries, illnesses, or circumstances considered highly suspicious for child maltreatment are easier to identify and to be accurately documented using ICD codes, such as subdural hemorrhage, skull fractures, and certain household circumstances that are observed in children under specific ages (Schnitzer et al., 2011). Given the likely underestimation of the true magnitude of child maltreatment when exclusively depending on ICD codes for maltreatment, it is imperative to develop a set of criteria for selecting medical conditions that are reliably related to suspected maltreatment. While this approach is probabilistic rather than deterministic, it offers an opportunity to enhance the utility of ICD codes in clinical and research settings and facilitate the comparison of the prevalence and trends of suspected child maltreatment across studies (Scott et al., 2009).

We completed, to our knowledge, the first scoping review to summarize how studies have selected and used ICD codes of injuries, illnesses, conditions, or circumstances that are suggestive of child maltreatment without the presence of a medical diagnosis of maltreatment. The objectives of this study were to 1) examine the types of research being conducted with ICD codes related to “suggestive” or “potential” maltreatment, 2) summarize the methods used to identify the ICD codes, and 3) propose future direction. This study could lay the foundation for future research into the operationalization of suspected/potential maltreatment in health care records.

## Methods

### Inclusion and Exclusion Criteria

For the purpose of this study, an ICD code suggestive of child maltreatment is a code of illness, injury, or circumstance that could indicate that a child has possibly or probably experienced maltreatment.

Studies included in the systematic review met the following eligibility criteria:

1. Published in peer-reviewed journals in English,
2. Included participants aged 0-18 years, and
3. Reported ICD codes suggestive of child maltreatment (i.e., physical abuse, neglect, medical neglect, sexual abuse, or emotional abuse) in any medical setting.

Studies published in conference proceedings, abstracts, or dissertations were omitted. Articles that did not mention the use of ICD codes, those that identified suspicious child maltreatment or injury types but did not mention that ICD codes were used to identify the cases, or those that only included explicit ICD codes for child maltreatment (e.g., ICD-9-CM code 995.5) were excluded.

### Search Strategy and Data Sources

This review adhered to the Preferred Reporting Items for Systematic Reviews and Meta-Analyses Extension for Scoping Reviews (PRISMA-ScR) guidelines (Tricco et al., 2018). A medical librarian (SJK) with expertise in systematic searching composed a sensitive search balancing subject headings and keywords to represent the concepts of International Classification of Diseases codes, children, and abuse or neglect. The databases MEDLINE via PubMed, Embase via Elsevier, Scopus via Elsevier, the Cumulative Index to Nursing and Allied Health Literature via Ebsco, and APA PsycINFO via Ebsco were searched from inception to September 1, 2020. The search was revised to include the concept of “non-accidental trauma” or “non-accidental injury” in the section capturing abuse and neglect and updated on April 16, 2021. All results were compiled in EndNote (The EndNote Team, 2013) and imported into Covidence (Covidence systematic review software, 2019) for deduplication and screening. Full details of the search strategies are available in Supplemental Table S1. The reference lists from the eligible articles in the full-text screen phase were also hand-searched to identify potential additional relevant studies.

### Study Selection and Data Extraction

Two reviewers (YL and BJ) independently screened the titles, abstracts, and the full texts identified using Covidence. Disagreements about study eligibility were resolved by achieving consensus through discussion or by a third reviewer (EG). Key information from the final identified full texts was extracted into Microsoft Excel by two reviewers (YL and BJ) and re-read and verified for accuracy and completeness by two reviewers (YL and JB). Extracted data included articles’ information (e.g., author, year, and journal), study characteristics (e.g., country, study design), child characteristics (e.g., age, sex, and race), data sources, study objectives, study settings, forms of child maltreatment (e.g., physical, emotional, sexual abuse, and neglect), ICD codes suggestive of maltreatment, excluded co-occurring ICD codes, and methods utilized to identify ICD codes.

### Data Synthesis

We used a narrative approach to synthesize the key findings regarding how each study identified and used ICD codes suggestive child maltreatment. Studies were grouped by maltreatment type (i.e., physical abuse only, abusive head trauma only, sexual abuse only, and multiple types of maltreatment). Then within each maltreatment type, we summarized characteristics of the study, the methods used to identify ICD codes suggestive of the study, and whether or not the study incorporated co-occurring exclusionary ICD codes. Due to the heterogeneity of the goals of the identified studies, it was beyond the scope of this study to quantitatively synthesize findings.

## Results

### Study Characteristics

Our search generated 3,638 articles (Figure 1). After removing duplicates, 1,135 titles and abstracts were screened and 90 met the inclusion criteria. The full-text of these 90 articles were assessed and 55 were excluded for the following reasons: 25 only included explicit ICD codes for child maltreatment, 16 were not peer-reviewed journal articles, nine did not use ICD codes, two were duplicates, two were not in English, and one focused on adults. By reviewing the reference lists of included studies, we identified two additional articles, leading to a final sample of 37 studies in 26 journals. Tables 1 provides a description of the characteristics of included studies.

**Figure 1.**
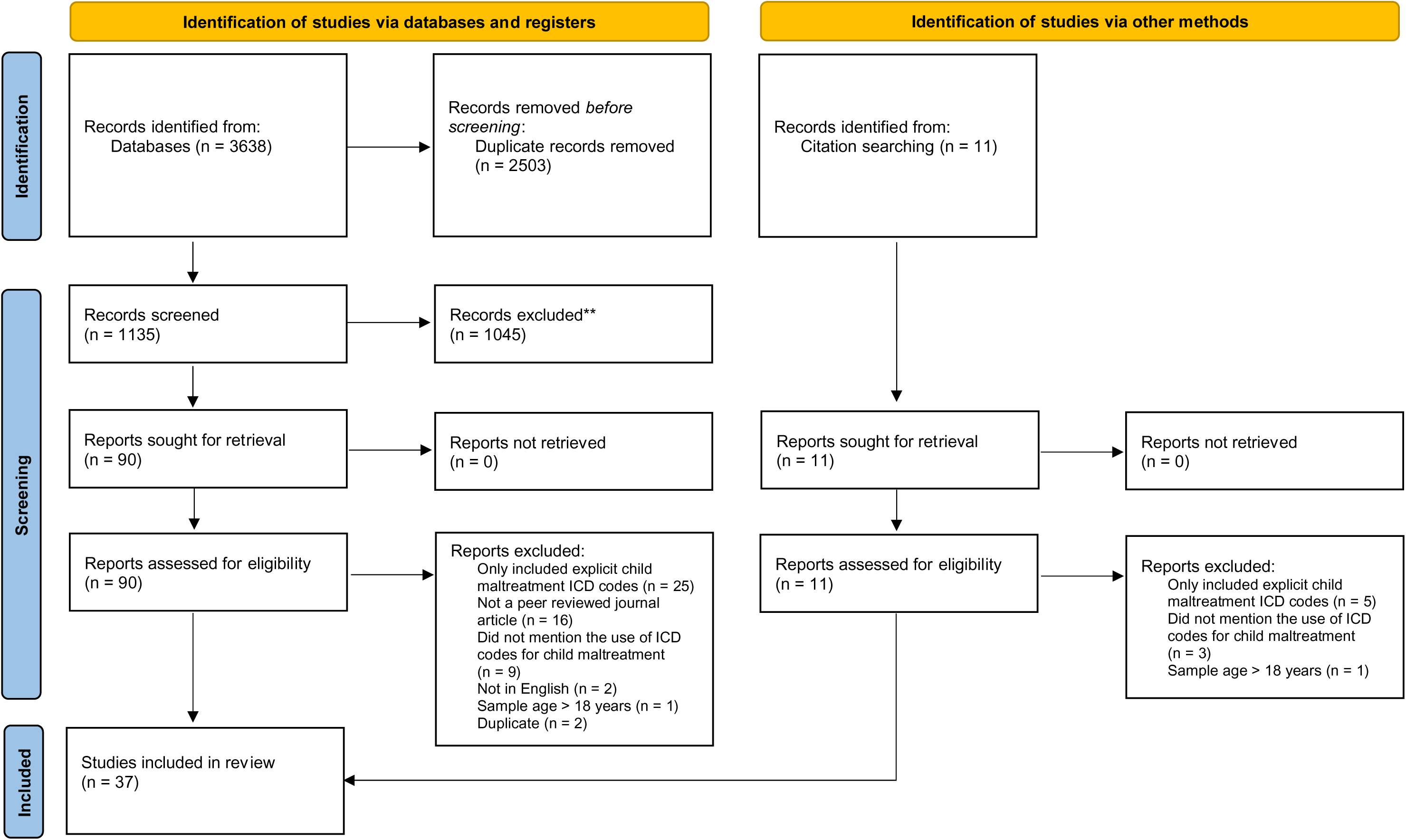
PRISMA flow diagram of the 37 selected studies.

**Table 1.**
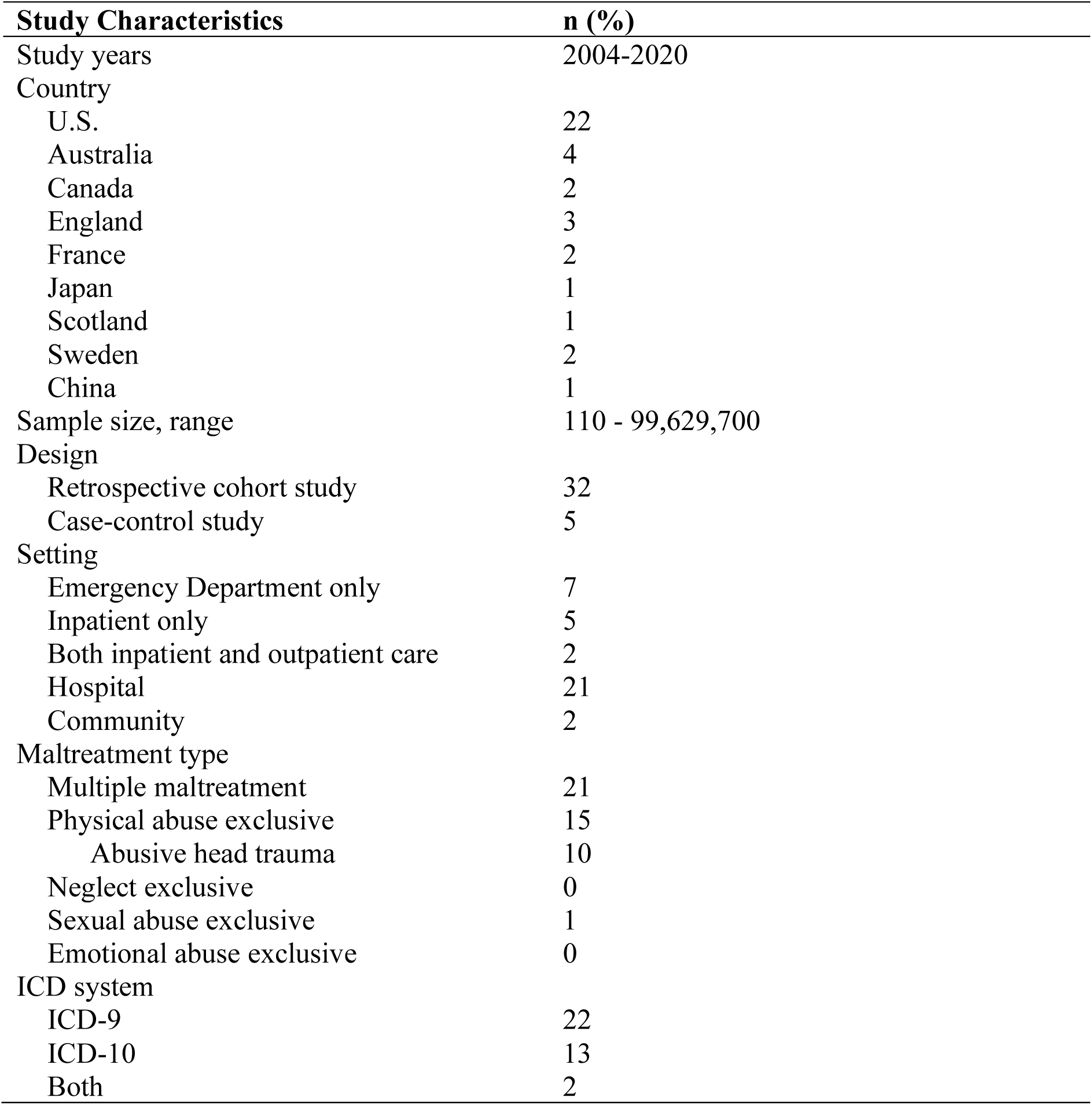
Description of 37 Included Studies

| **Study Characteristics** | **n (%)** |
| --- | --- |
| Study years | 2004-2020 |
| Country |  |
| U.S. | 22 |
| Australia | 4 |
| Canada | 2 |
| England | 3 |
| France | 2 |
| Japan | 1 |
| Scotland | 1 |
| Sweden | 2 |
| China | 1 |
| Sample size, range | 110 - 99,629,700 |
| Design |  |
| Retrospective cohort study | 32 |
| Case-control study | 5 |
| Setting |  |
| Emergency Department only | 7 |
| Inpatient only | 5 |
| Both inpatient and outpatient care | 2 |
| Hospital | 21 |
| Community | 2 |
| Maltreatment type |  |
| Multiple maltreatment | 21 |
| Physical abuse exclusive | 15 |
| Abusive head trauma | 10 |
| Neglect exclusive | 0 |
| Sexual abuse exclusive | 1 |
| Emotional abuse exclusive | 0 |
| ICD system |  |
| ICD-9 | 22 |
| ICD-10 | 13 |
| Both | 2 |

Over time, the number of studies using ICD codes suggestive of maltreatment has increased, with three published between 2000 and 2010, and 34 between 2011 and 2021. Most studies relied on U.S. samples (n = 22, 59.5%), followed by Australia (n = 4, 10.8%), England (n = 3, 8.1%), Canada, France, and Sweden (n = 2 each, 5.4%), Japan, Scotland, and China (n = 1 each, 2.7%). Most of the included studies (n = 32, 86.5%) were non-comparator retrospective cohort studies using administrative data to ascertain the prevalence of child maltreatment. The age of study participants varied. Twelve studies included children under 19 years, six under 11 years, seven under six years, 11 under four years, and only one under one year. Fifteen studies exclusively focused on physical abuse, among which 10 examined abusive head trauma (AHT); one study focused on sexual abuse, while the rest examined multiple types of maltreatment. The majority of studies used ICD-9 coding system (n = 22, 59.5%) compared to the ICD-10 system (n = 13, 35.1%), and two studies (5.4%) used both coding systems. Most of the studies (n = 36, 97.3%) reported the entire code set used. Two studies (5.4%) used disease manifestation codes only, one study (2.7%) used the external injury cause codes only, and most of the studies (n = 34, 91.9%) used a combination of both types of codes. The complete list of ICD codes can be provided upon request.

### The Purpose of the Study and Coding Scheme by Maltreatment Type

Table 2 presents the key study characteristics, including the main study aim, country, data source, sample size, age range, methods identifying the ICD codes, and the coding system used.

**Table 2.**
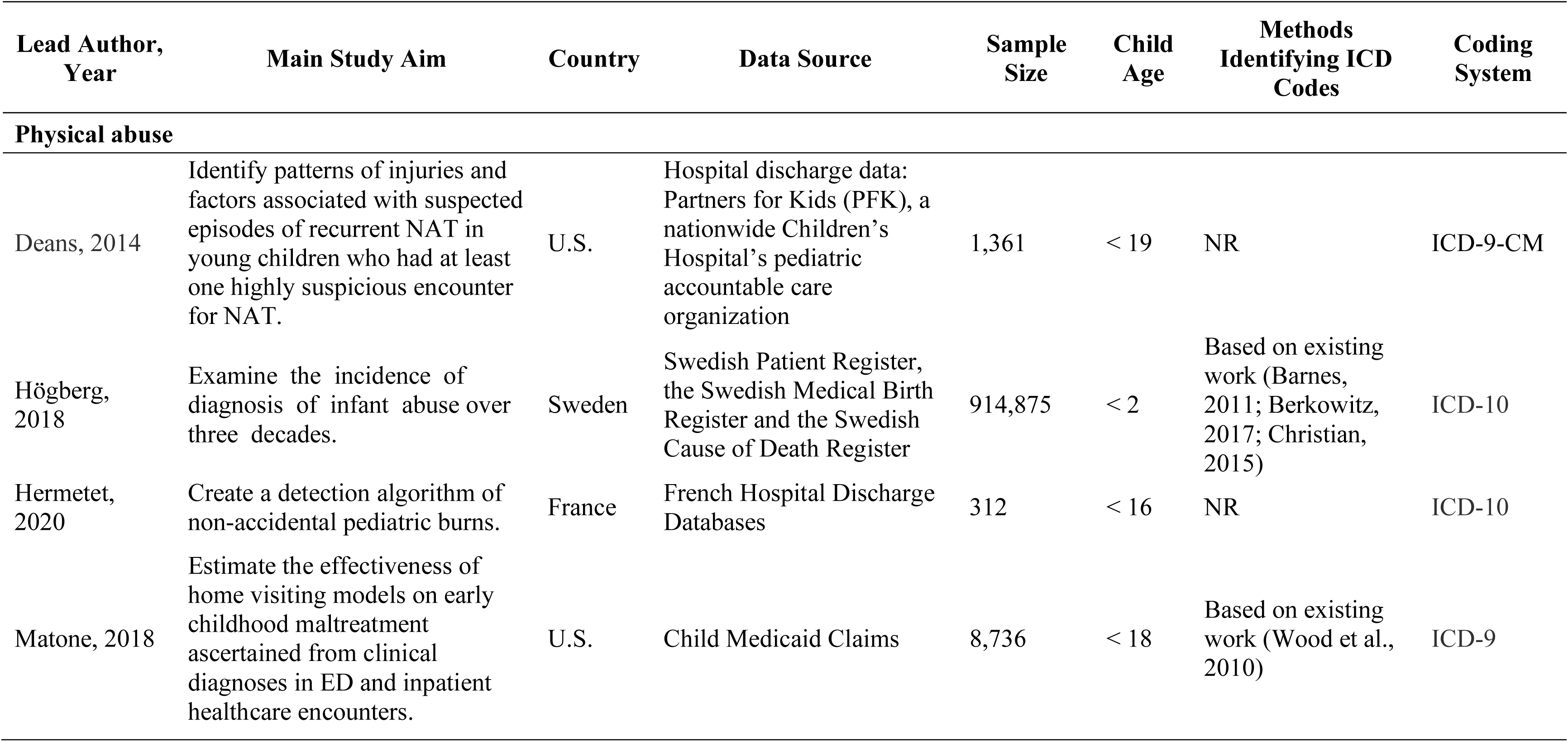
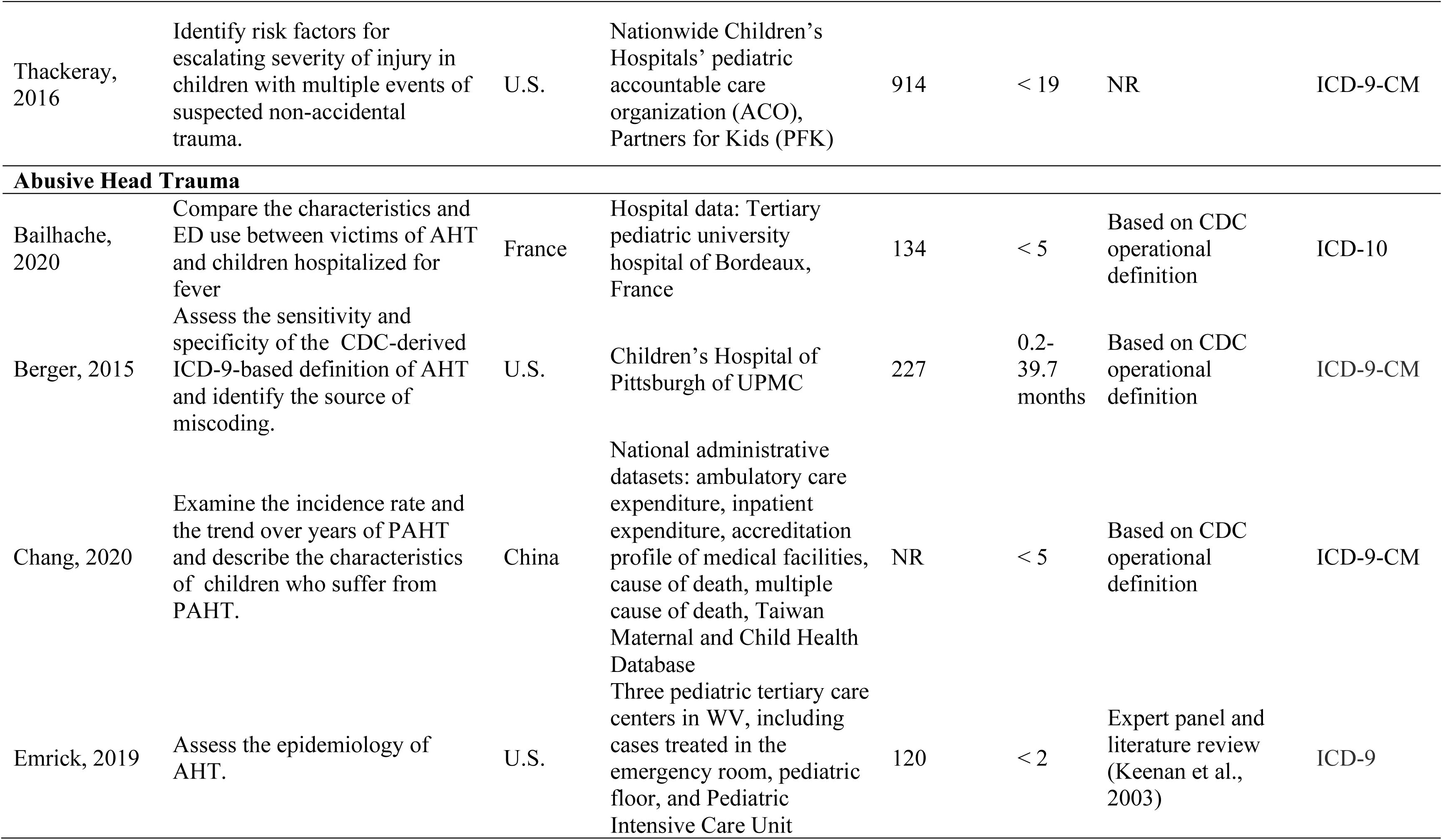
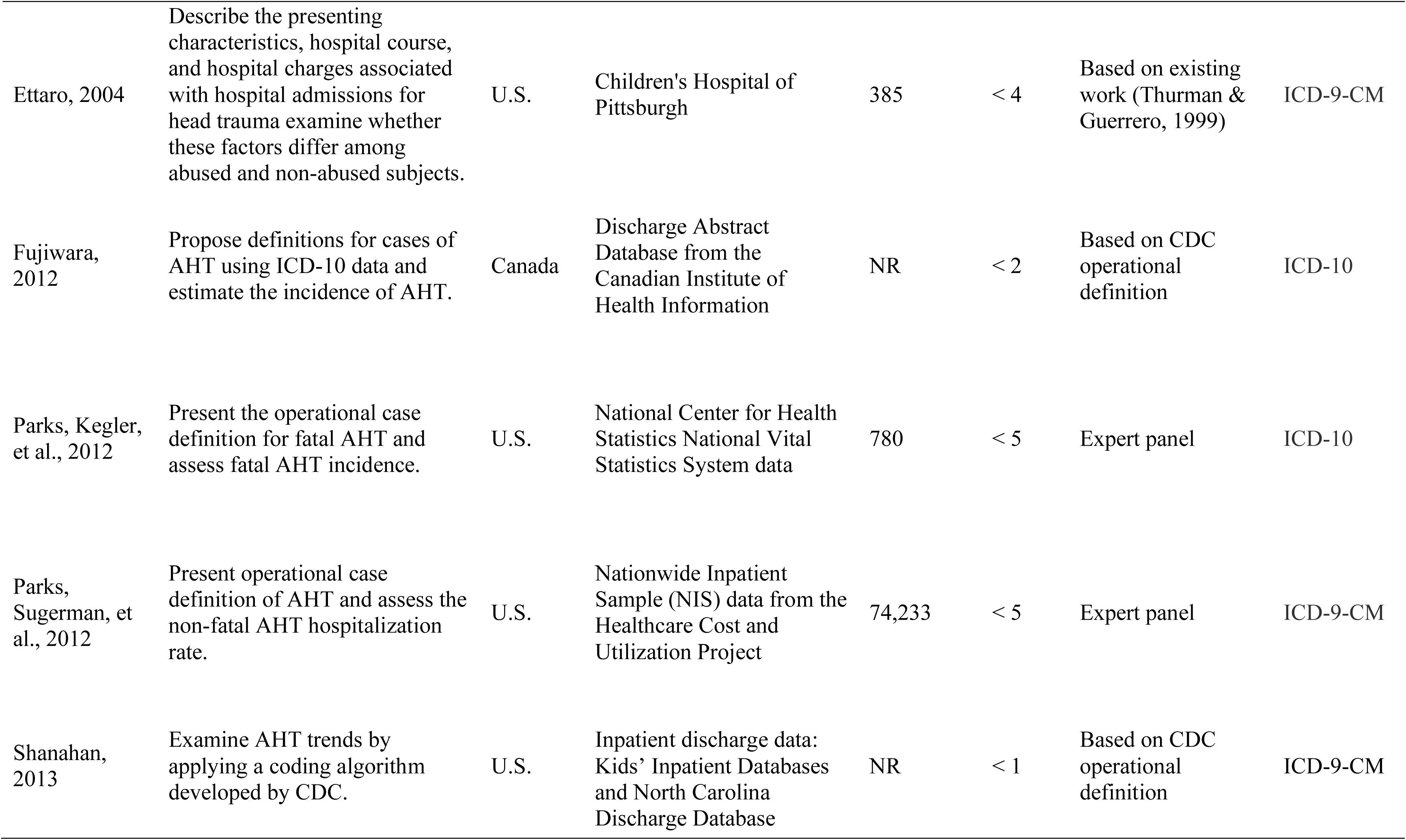
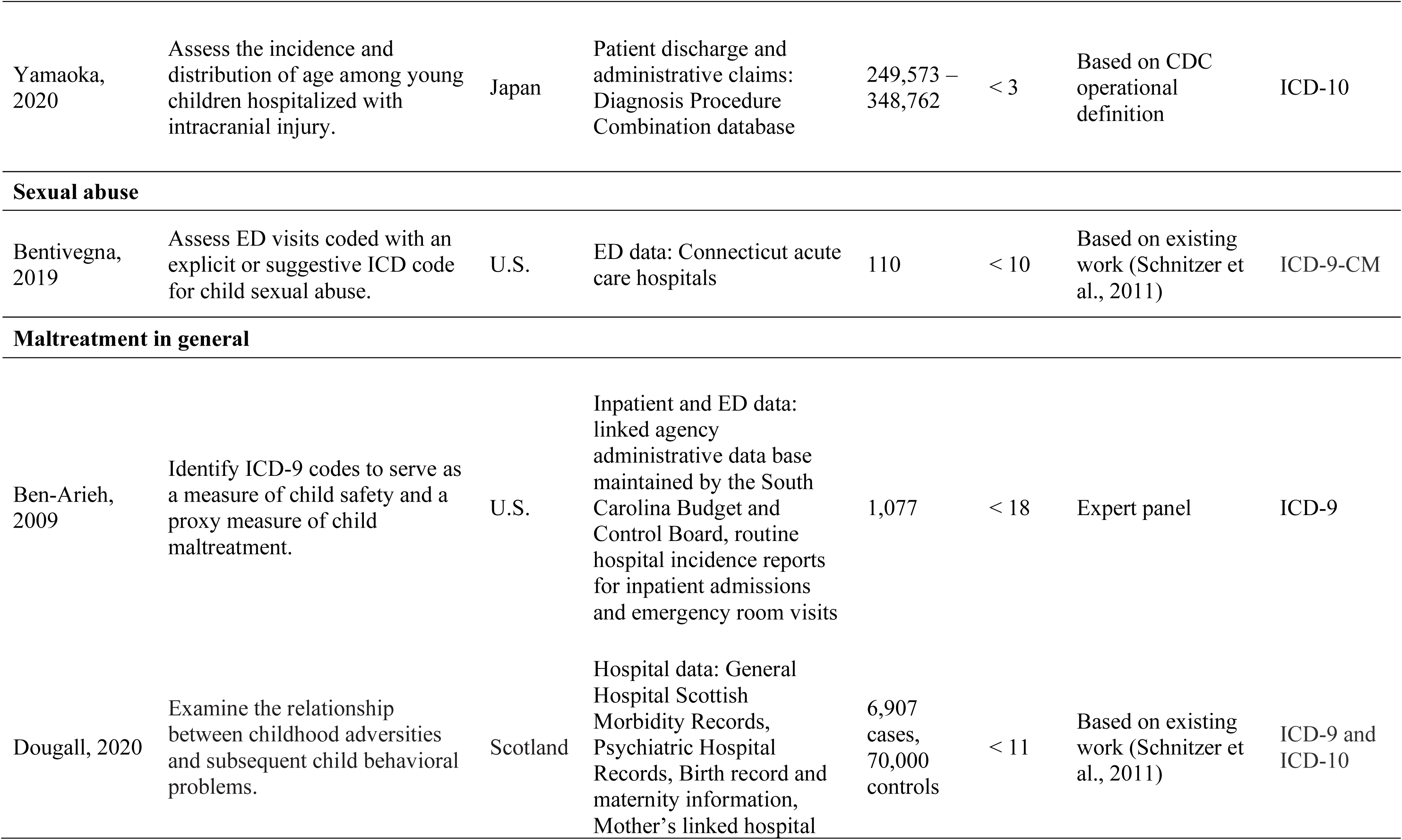
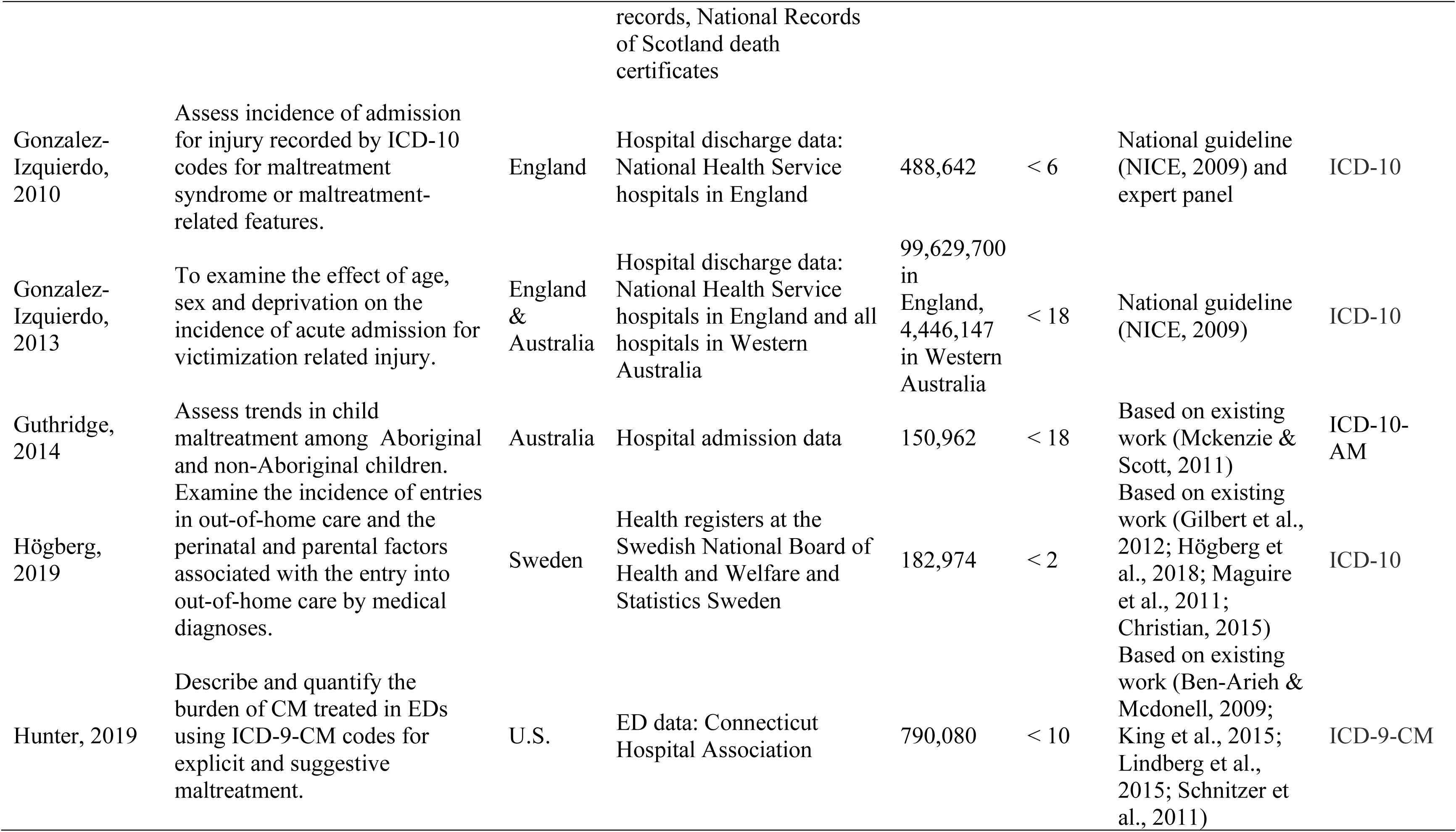
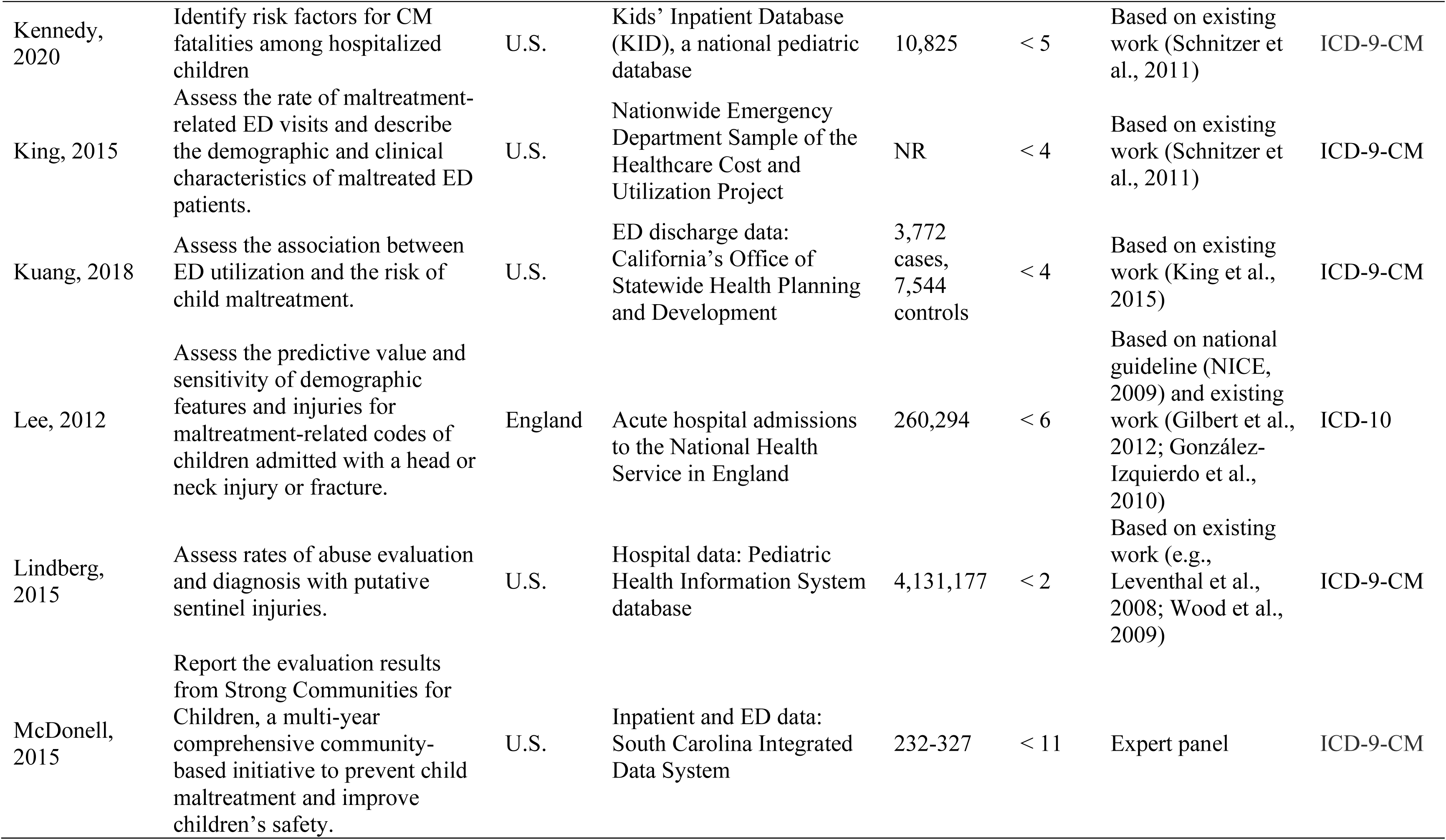
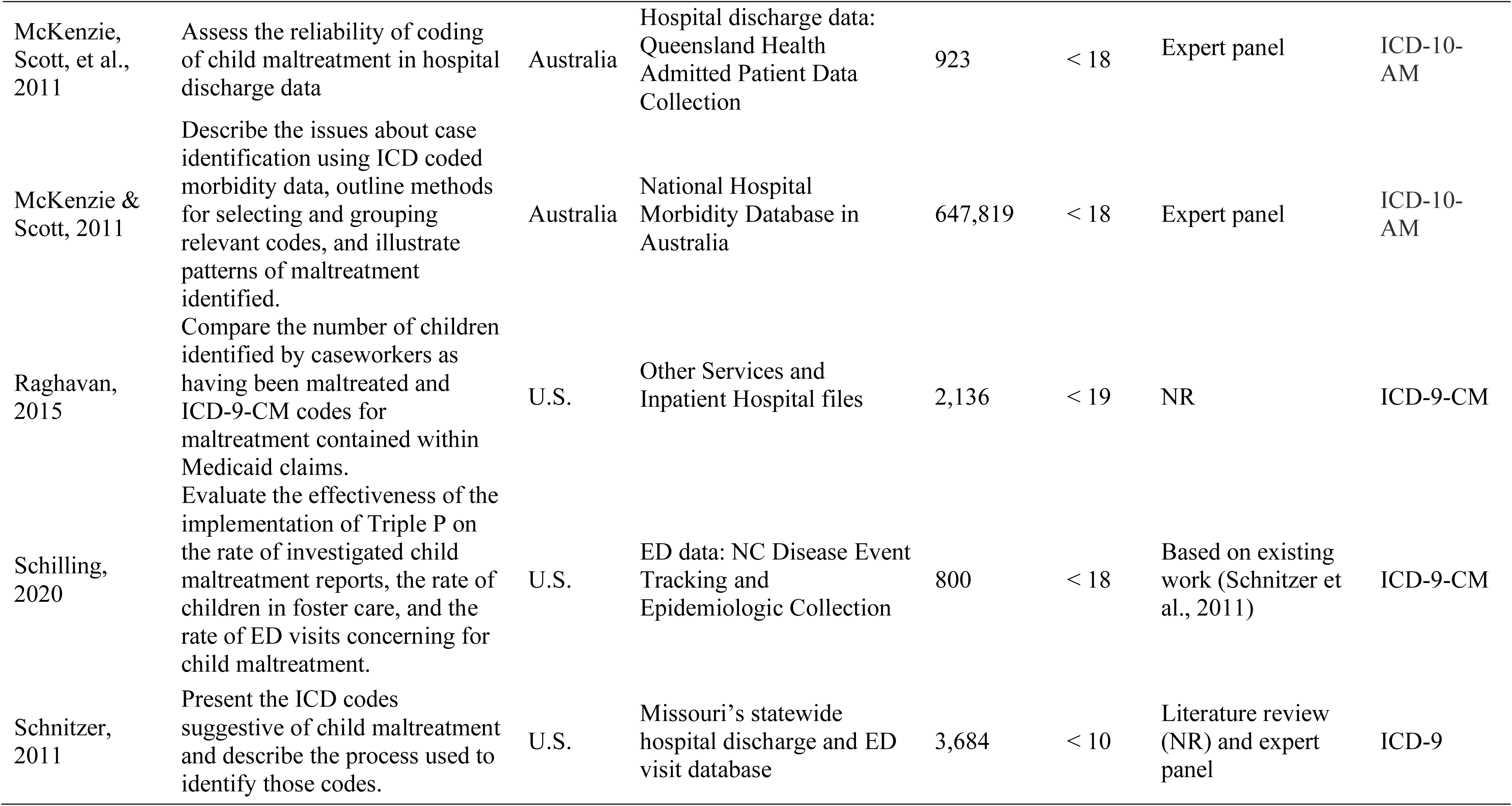
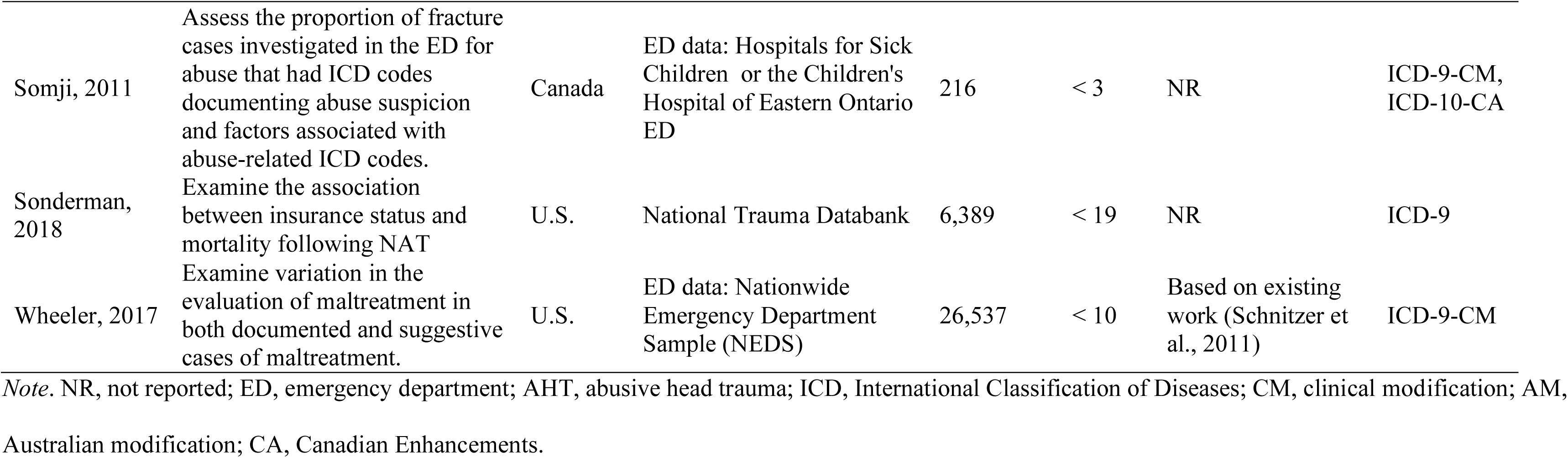
Selected Study Characteristics of Included Studies

### Multiple Maltreatment

Twenty-one studies simultaneously examined multiple types of maltreatment, with 18 focusing on physical abuse, neglect, and sexual abuse, and three also including emotional or psychological abuse (Guthridge et al., 2014; McKenzie et al., 2011; McKenzie & Scott, 2011). Among the included studies which focused on multiple maltreatment, the most common research objective (11 studies) was to determine the incidence or the trends of maltreatment-related injuries (Ben-Arieh & McDonell, 2009; Gonzalez-Izquierdo et al., 2010; Guthridge et al., 2014; Hunter & Bernstein, 2019; King et al., 2015; Lee et al., 2012; Lindberg et al., 2015; McDonell et al., 2015; Schilling et al., 2020; Somji et al., 2011; Wheeler et al., 2017). Four articles described the demographic and clinical characteristics of patients suspected for maltreatment (King et al., 2015; Kuang et al., 2018; Somji et al., 2011; Sonderman et al., 2018), two studies validated the ICD codes for maltreatment related injuries or circumstances (McKenzie et al., 2011; Raghavan et al., 2014), three focused on the process of identifying a set of ICD codes suggestive of maltreatment (Ben-Arieh & McDonell, 2009; McKenzie & Scott, 2011; Schnitzer et al., 2011), and three examined the relationships between maltreatment related injuries and child outcomes (e.g., child maltreatment fatalities; mental health, suicidal behavior, and out-of-home care; Dougall et al., 2020; Högberg et al., 2019; Kennedy et al., 2020).

The coding scheme of suggestive maltreatment based on precedent from Schnitzer et al. (2011) was applied by seven articles (Dougall et al., 2020; Hunter & Bernstein, 2019; Kennedy et al., 2020; King et al., 2015; Kuang et al., 2018; Schilling et al., 2020; Wheeler et al., 2017), making it a seminal study in the research on ICD codes for suspected maltreatment. This study first compiled a comprehensive list of ICD codes suspicious for child maltreatment using literature review and consultation with an advisory panel of experts on child maltreatment. The authors then selected a sample of 3,634 hospital and ED visits by children under 10 years old who were diagnosed with suspicious maltreatment. The abstracted medical records of these visits were reviewed by the project team and an advisory panel, and an ICD code was determined to be suggestive of maltreatment if more than 66% of the visits with the diagnostic code indicated probable or possible maltreatment. This study identified 37 ICD-9-CM codes with different usage rules by age and incorporated exclusionary criteria for co-occurring ICD codes.

A coding scheme proposed by McKenzie & Scott (2011) was applied by two studies (Guthridge et al., 2014; McKenzie et al., 2011). This study selected maltreatment-related codes by systematically reviewing the ICD-10 Australian Modification (ICD-10-AM) classification system and consultation with expert health information managers. Although the codes identified by this study mostly included those specific for maltreatment syndromes, it also included codes that were rarely seen in other studies for suspected neglect (e.g., problem related to upbringing) and emotional or psychological abuse (e.g., hostility towards and scapegoating of child, inappropriate parental pressure and other abnormal qualities of upbringing). Two studies (Ben-Arieh & McDonell, 2009; McDonell et al., 2015) proposed ICD codes for child safety as a proxy for child maltreatment as determined by an expert panel. The coding scheme developed by one of the studies (Ben-Arieh & McDonell, 2009) identified 323 codes suggestive of child physical and sexual safety hazards, and child safety not otherwise classified. Each type of safety hazard categories contained sub-classifications, such as fracture of skull, viral diseases accompanied by exanthem, or diseases of other endocrine glands; however, the full list of codes was not provided.

The United Kingdom National Institute of Health and Clinical Excellence (NICE) clinical guidelines were used by two England-based studies (Gonzalez-Izquierdo, 2010; Lee, 2012) to develop measures of suspected child maltreatment using ICD-10 codes (National Institute of Health and Clinical Excellence, 2009). The NICE guideline includes recommendations on physical features (e.g., bruises, bites, and intracranial injuries); clinical presentations (e.g., pregnancy, poisoning, inappropriately explained poor school attendance); failure of provision and failure of supervision; emotional, behavioral, interpersonal and social functioning; and parent-child interactions. These features should prompt the clinician to suspect child maltreatment and consider further action. Studies following NICE identified the following injuries as associated with a high risk of maltreatment: head and neck injuries, intracranial injury, head injury (no fracture), and fractures (e.g., skull fracture, long bone fracture, thoracic fracture, and fracture of facial bones) (National Institute of Health and Clinical Excellence, 2009). A list of codes for cause of injury was also included and classified based on a hierarchy that indicate likelihood of maltreatment. For example, assault or undetermined cause reflect a higher likelihood of maltreatment, while adverse social circumstance reflects a diminishing likelihood.

### Physical Abuse

Five articles exclusively focused on physical abuse (age range: 2 - 18 years; Deans et al., 2014; Hermetet et al., 2020; Högberg, 2018; Matone et al., 2018; Thackeray et al., 2016). Reported outcomes of the four articles were all about the prevalence or rate of physical abuse related injuries. Among the four articles, one exclusively focused on non-accidental pediatric burns (NAB) in France (Hermetet et al., 2020) to build a detection algorithm of NAB. Hermetet et al. (2020) selected ICD-10 codes for intentional violence, events of undetermined intent, or possible consequences of maltreatment to classify children as a suspect of maltreatment. Matone et al. (2018)’s article estimated the effectiveness of a home visiting model on early childhood maltreatment and included injuries mostly about fractures (e.g., fractures of the femur, radius, ulna, tibia, fibula, humerus, ribs). This study also identified traumatic brain injuries as suggestive of abuse for children under 2 years. Thackeray et al. (2016) and Dean et al. (2014)’s studies used a broad range of ICD codes suspicious for abuse (e.g., ICD-9-CM codes 800-904, 910-957) to indicate a diagnosis of non-accidental trauma (NAT) and examined risk factors for children with multiple events of suspected NAT; however, the descriptions of the codes were not provided.

### Abusive Head Trauma

Although AHT is a subtype of physical abuse, we presented the studies separately from physical abuse because most of the ICD codes used for AHT differed from those used for physical abuse. Overall, 10 articles focused on AHT, with children’s ages all under five years (age range: 0.2 months - 4 years). Most of the studies (n = 7) estimated the incidence rate of AHT (Chang et al., 2020; Emrick et al., 2019; Fujiwara et al., 2012; Parks, Kegler, et al., 2012; Parks, Sugerman, et al., 2012; Shanahan et al., 2013; Yamaoka et al., 2020), two studies compared the demographic characteristics or health care utilization between victims of AHT and other children (Bailhache et al., 2020; Ettaro et al., 2004), two focused on describing the characteristics of children with AHT (Chang et al., 2020; Yamaoka et al., 2020), and one assessed the sensitivity and specificity of AHT coding recommended by the Centers for Disease Control and Prevention (CDC) (Berger et al., 2015). Among the 10 articles, seven (Bailhache et al., 2020; Berger et al., 2015; Chang et al., 2020; Fujiwara et al., 2012; Parks, Kegler, et al., 2012; Parks, Sugerman, et al., 2012; Shanahan et al., 2013) used the definition and codes for AHT recommended by CDC (Parks, Annest, et al., 2012; excluded from our review because it is not a peer-reviewed article). The CDC developed a list of ICD-9-CM codes for non-fatal AHT with a panel of experts on child maltreatment. The codes were identified for broad and narrow definitions of AHT in children less than five years old, with the former focusing on increased sensitivity and the latter focusing on increased specificity. Each definition included two sets of ICD codes indicating definitive or presumptive AHT and probable AHT. To be classified as presumptive or probable AHT, it requires the presence of an ICD code and an assault-related E code. Two articles (Emrick et al., 2019; Yamaoka et al., 2020) coupled the CDC recommendation with elements from previous studies to define AHT for children under two and three years, and both studies included retinal hemorrhage (362.81) in addition to the CDC recommended codes. Ettaro et al. (2004) selected codes based on a previous study that identified traumatic brain injury events (Thurman & Guerrero, 1999) but most of the included codes were mirrored those from the CDC’s recommendation.

### Sexual Abuse

Only one article exclusively focused on sexual abuse. Bentivegna et al. (2019)’s study compared the prevalence of child sexual abuse and the demographic characteristics of victims of sexual abuse by different coding types (i.e., ICD codes for explicit and suggestive sexual abuse). They adopted codes for child sexual abuse from Schnitzer et al. (2011)’s study for children under 10 years old and included codes for genital herpes, gonococcal infection, pelvic inflammatory disease, contusion of genital organs, and observations after alleged rape or seduction. The code for “observation for abuse/neglect” (V71.81) in Schnitzer et al. (2011)’s article was excluded from their coding list.

### Validation of ICD Codes

Of all the included studies, only four estimated the sensitivity and specificity of the ICD codes (Berger et al., 2015; Hermetet et al., 2020; McKenzie et al., 2011; Raghavan et al., 2015). Three studies validated a mixture of codes for specified and suggestive maltreatment simultaneously, thus it was not possible to distinguish whether the levels of ascertainment were due to the specified codes or the suggestive codes. Specifically, Berger et al. (2015) validated the CDC-derived ICD codes for AHT using the evaluation by a Child Protection Team at a tertiary care children’s hospital, and McKenzie et al. (2011) ascertained the codes for maltreatment by conducting a retrospective medical record review. Both studies demonstrated high sensitivity (92% - 98%) and specificity (95% - 96%). Using assessment of maltreatment by caseworkers at Child Protective Services agencies as a gold standard, Raghavan et al. (2015)’s study found that only 15% of children involved in child welfare agencies were identified as maltreatment by ICD codes and 16% were false-positive cases. Hermetet et al. (2020) developed an algorithm for the detection of maltreatment by non-accidental burns and performed a medical chart review, and found that the sensitivity was 90% for the narrow definition of maltreatment, which excluded child neglect, and 48% for the broad definition of maltreatment, while specificity was between 65% to 70%.

### Excluded Co-Occurring ICD Codes

The majority of studies assessing ICD codes suggestive of child maltreatment actively excluded certain co-occurring ICD codes. The most frequently excluded codes were those of transport accidents (n = 15), birth trauma (n = 8), bleeding disorders (n = 4), injuries due to fall (n = 4), coagulation defects (n = 3), osteogenesis imperfecta (n = 3), and prematurity (n = 3). Other excluded codes included accidents due to natural and environmental events, adverse effects of drugs, medicinal, and biological substances in therapeutic use, suicide and self-inflicted injury, legal intervention (e.g., apprehension by the police), injuries occurring at an industrial place or premises, or at a place for recreation and sport, osteochondrodysplasia, congenital anomalies, unintentional gun-related injuries, purpura, other hemorrhagic conditions, and deficiency of vitamin K. Eighteen studies did not report the use of excluded co-occurring codes. The complete list of excluded co-occurring codes can be provided upon request.

### Methods Used to Identify ICD Codes

Literature review is the most commonly used method to identify ICD codes suggestive of child maltreatment (n = 23). As mentioned above, two seminal studies (McKenzie & Scott, 2011; Schnitzer et al., 2011) and two guidelines (Parks, Annest, et al., 2012; NICE, 2009) were frequently cited by other articles. Six studies identified the ICD codes by consulting with expert panels to classify the injuries or illness as maltreatment related or not. Two studies used both expert panel and literature review to identify codes. However, six studies did not provide information on how the ICD codes were selected.

## Discussion

Our study is the first scoping review that systematically explored how ICD codes were used in research to identify possible or suspected maltreatment in administrative health services data. Thirty-seven articles were included and over half of the articles examined multiple types of child maltreatment. Growth in the number of studies relying on ICD codes to document children exposed to events which were plausibly related to maltreatment suggests a heightened urgency in understanding the validity, accuracy, and limitations of such measures. Our results demonstrate that some injury or illness patterns may signal suspected maltreatment. However, studies varied greatly in the types of codes used, whether or not to employ exclusionary condition, how they identified relevant codes, and the developmental ages when specific codes were relevant. This heterogeneity coupled with lack of clear rationale for such variation may have unintended consequences such as lack of comparability across studies and lack of clear guidance for future research. Nevertheless, we hope that by providing access to a summarized list of ICD codes and methods used to develop those codes, future researchers will be able to critically choose measures of suspected maltreatment from the list we provided or develop their own set of codes based on the existing ones without reinventing the wheel.

A broad range of ICD codes representing a diverse range of diagnosed health conditions (e.g., burns, fractures, presumed reasons for injury) were used to identify possible or suspected maltreatment in the included studies. Researchers identified ICD codes suggestive of maltreatment reliably using administrative health data through literature reviews and expert panels. Despite these systematic approaches, researchers drew different conclusions regarding what codes to use, resulting in substantial variability in coding, age range for codes, and exclusion criteria for co-occurring conditions. The purpose of most of the studies reviewed here was to estimate the incidence of maltreatment. However, substantial heterogeneity in which codes are applied for inclusion or exclusion likely affect the reported incidence. Thus, the reported incidences of child maltreatment are not comparable across studies.

Complicating the selection of ICD codes, is how these codes vary with age. From the full text of studies, it was difficult to assess whether the ICD codes for an age group were chosen because of the development applicability of the codes or because of the idiosyncratic nature of the sample. This distinction is important for understanding how to apply the existing research in future contexts. For instance, Schnitzer et al. (2011)’s study included children through younger than 10 years because their funding program excluded older children; however, it was unclear which of their codes may apply to older youth. In fact, the same code set was applied by another study of children under 18 years old without justification (Schilling, 2020). Likewise, rib fracture was identified as suggestive of physical abuse for children under two (Högberg, 2018; Lindberg, 2015), five (Schnitzer et al., 2011), and 18 (Matone, 2018; Schilling, 2020), while a meta-analysis on fracture in young children and infants and child maltreatment found that children under the age of three presenting with a rib fracture should raise the awareness for a child abuse evaluation (Mitchell et al., 2021). Because relevant codes for suspected maltreatment vary with age, future research proposing coding schemes should explicitly discuss upper and lower age limits for each diagnosis code.

Exclusion criteria for co-occurring diagnoses offer a means to increase certainty that the codes applied were due to maltreatment rather than other causes (i.e., an accidental rather than non-accidental event). Many included studies did not report incorporating exclusion of co-occurring codes. Lack of use of exclusion criteria would likely lead to the prevalence of suspected maltreatment being overestimated. However, in practice, co-occurring exclusionary codes are challenging to implement. For example, the exclusionary code may be documented in a separate but related encounter as the diagnosis of injury or illness suggestive of maltreatment. Future efforts should provide detailed information or technical instructions on how exactly to apply such exclusion criteria.

To understand the scope of research questions for which a given coding scheme is appropriate, it is essential to evaluate these coding schemes; however, only four of the included studies validated the codes they identified and most of them performed the validation for specified and suggestive maltreatment codes simultaneously. Most of the included studies applied code sets previously used without further assessing the coding schemes’ sensitivity and specificity in their specific health care settings and samples. Yet, the diagnostic performance of the ICD codes may change in different clinical settings (Hohl et al., 2014). Thus, more studies assessing the sensitivity and specificity of case ascertainment in administrative health data to identify suspected child maltreatment are needed. A promising approach is to use patient-level data integrated with information from other sources such as formal assessments conducted by specialized child abuse pediatricians. Further, the assessments should be examined separately by setting (e.g., hospital vs. ambulatory care), clinician characteristics (e.g., specialty, experience, etc.) and patient characteristics (e.g., age, sex, and race/ethnicity) (McKenzie & Scott, 2011).

To date, CDC has published two guidelines regarding specified child maltreatment (Leeb et al., 2008) and AHT (Parks, Annest, et al., 2012), both were intended to build a uniform definition and consistency for the application of these definitions to monitor the incidence and changes of child maltreatment over time, identify children at highest risk of maltreatment for early prevention, and assess the effectiveness of prevention and intervention programs (Leeb et al., 2008; Parks, Annest, et al., 2012; Saltzman et al., 1999). Our review found that the ICD codes for AHT were most consistently applied in the included studies. This consistency was largely contributed to the guidance provided by CDC’s definition and recommendation. The guidelines provided uniform definitions, recommended data elements, or recommended sets of ICD diagnosis codes and external cause of injury codes for definite or probable AHT. Future efforts to established standardized operational definitions and common code sets for suspected child maltreatment may be based on the approach used for these reports.

There are several key limitations of the present review that warrant discussion. First, language bias may affect the generalizability of current findings (Stern & Kleijnen, 2020). Only English language publications were included. The lack of evidence in countries other than the U.S., Australia, and European countries may be partly due to language restriction. Furthermore, we excluded meeting abstracts or conference proceedings; however, such publications do not always provide enough information, such as the set of codes used to fulfil our research objectives. Third, the variability in the coding systems (ICD-9 vs. ICD-10) applied and national coding versions (e.g., ICD-10-AM) created additional complexity. Additionally, we did not include articles that only used ICD-10 codes for suspected maltreatment (e.g., T76.1 for physical abuse, suspected). However, it is worth noting that these codes are also under-used (Durand, 2019; Hunter et al., 2021) and thus inadequately captured the breadth of suspected maltreatment. Instead, we focused on codes where the accidental or non-accidental nature of the event leading to the condition was ambiguous. Given the limitations of existing studies, our review stopped short of recommending a set of ICD codes for operationalizing a definition of suspected maltreatment. Nevertheless, our review yielded an extensive list of ICD codes to serve as the foundation for future investigation and consensus building.

Both over and under identification of child maltreatment can have negative consequences for children and families. Health care providers have a unique vantage points into children’s lives and thus their administrative records hold great promise for studying child maltreatment at the population level. The World Health Organization (WHO) in collaboration with the International Society for Prevention of Child Abuse and Neglect (ISPCAN) has called for common conceptual and operational definitions of child maltreatment and the set-up of working groups to develop an agreed set of guidelines on assigning ICD codes to confirmed and suspected child maltreatment cases (World Health Organization, 2006). Our review provides a basis for developing recommended guidelines that establish a comprehensive and uniform set of codes for studying child maltreatment. The establishment of clear guidelines and the validation of the codes could enable researchers to more efficiently and uniformly address a variety of questions such as estimating prevalence and cost of care, and evaluating prevention efforts.

## Data Availability

All data produced in the present work are contained in the manuscript

## Acknowledgements

We would like to thank Kelly Evans, MPH (Center for Child and Family Policy, Duke University) for inspiring us to pursue this research inquiry and provide feedback on the manuscript.

---

**Supplemental Table S1.** Search Strategy

| Set # |  | Results |
| --- | --- | --- |
| 1 ICD | "International Classification of Diseases"[Mesh] OR ((ICD[tiab] OR "International Classification of Diseases"[tiab]) AND (codes[tiab] OR code[tiab] OR coding[tiab] OR coded[tiab])) | 24625 |
| 2 Peds | ("Child"[Mesh] OR "Infant"[Mesh] OR "Adolescent"[Mesh] OR "Pediatrics"[Mesh] OR "Minors"[Mesh] OR "Puberty"[Mesh] OR infant[tiab] OR infants[tiab] OR infancy[tiab] OR newborn[tiab] OR newborns[tiab] OR neonatal[tiab] OR neonate[tiab] OR neonates[tiab] OR baby[tiab] OR babies[tiab] OR preterm[tiab] OR prematurity[tiab] OR toddler[tiab] OR toddlers[tiab] OR boy[tiab] OR boys[tiab] OR boyhood[tiab] OR girl[tiab] OR girls[tiab] OR girlhood[tiab] OR kid[tiab] OR kids[tiab] OR child[tiab] OR childhood[tiab] OR children[tiab] OR schoolchild[tiab] OR schoolgirl[tiab] OR schoolgirls[tiab] OR schoolboy[tiab] OR schoolboys[tiab] OR "school age"[tiab] OR "school aged"[tiab] OR preadolescent[tiab] OR preadolescents[tiab] OR preadolescence[tiab] OR adolescent[tiab] OR adolescents[tiab] OR adolescence[tiab] OR juvenile[tiab] OR juveniles[tiab] OR youth[tiab] OR youths[tiab] OR teen[tiab] OR teens[tiab] OR teenager[tiab] OR teenagers[tiab] OR teenaged[tiab] OR puberty[tiab] OR pubescent[tiab] OR pubescence[tiab] OR prepubescent[tiab] OR prepubescence[tiab] OR pediatric[tiab] OR pediatrics[tiab] OR paediatric[tiab] OR paediatrics[tiab] OR minors[tiab]) | 4,439,328 |
| 3 abuse | "Child Abuse"[Mesh] OR abuse[tiab] OR abused[tiab] OR abusive[tiab] OR abusing[tiab] OR abuser[tiab] OR abusers[tiab] OR maltreatment[tiab] OR maltreated[tiab] OR neglect[tiab] OR neglected[tiab] OR neglecting[tiab] OR neglects[tiab] OR "non accidental injury"[tiab] OR "non accidental injuries"[tiab] OR "non accidental trauma"[tiab] OR "non accidental traumas"[tiab] OR "nonaccidental injury"[tiab] OR "nonaccidental injuries"[tiab] OR "nonaccidental trauma"[tiab] OR "nonaccidental traumas"[tiab] OR "non-accidental injury"[tiab] OR "non-accidental injuries"[tiab] OR "non-accidental trauma"[tiab] OR "non-accidental traumas"[tiab] | 214,391 |
| 4 | 1 AND 2 AND 3 | 332 |
|  | 18822431 OR 10442662 OR 26142071 OR 31043070 OR 25631298 OR<br>22311999 OR 26438705 OR 21316104 | 7/8 |

Database (including vendor/platform): Embase
| Set # |  | Results |
| --- | --- | --- |
| 1 ICD | 'International Classification of Diseases'/exp OR ((ICD:ti,ab OR "International Classification of Diseases":ti,ab) AND (codes:ti,ab OR code:ti,ab OR coding:ti,ab OR coded:ti,ab)) | 73009 |
| 2 Peds | 'pediatrics'/exp OR 'infant'/exp OR 'child'/exp OR 'adolescent'/exp OR 'minor (person)'/exp OR 'puberty'/exp OR pediatric:ab,ti OR pediatrics:ab,ti OR paediatric:ab,ti OR paediatrics:ab,ti OR infant:ab,ti OR infants:ab,ti OR infantile:ab,ti OR baby:ab,ti OR babies:ab,ti OR preterm:ab,ti OR prematurity:ab,ti OR child:ab,ti OR children:ab,ti OR childhood:ab,ti OR toddler:ab,ti OR toddlers:ab,ti OR boy:ab,ti OR boys:ab,ti OR boyhood:ab,ti OR girl:ab,ti OR girls:ab,ti OR girlhood:ab,ti OR schoolchild:ab,ti OR schoolgirl:ab,ti OR schoolgirls:ab,ti OR schoolboy:ab,ti OR schoolboys:ab,ti OR 'school age':ab,ti OR preadolescent:ab,ti OR preadolescents:ab,ti OR preadolescence:ab,ti OR adolescent:ab,ti OR adolescents:ab,ti OR adolescence:ab,ti OR juvenile:ab,ti OR juveniles:ab,ti OR youth:ab,ti OR youths:ab,ti OR teenager:ab,ti OR teenagers:ab,ti OR teenaged:ab,ti OR teen:ab,ti OR teens:ab,ti OR pubescent:ab,ti OR 'young people':ab,ti OR 'young person':ab,ti OR 'young persons':ab,ti OR puberty:ab,ti OR pubescence:ab,ti OR prepubescent:ab,ti OR prepubescence:ab,ti OR minors:ab,ti | 4,863,617 |
| 3 abuse | 'child abuse'/exp OR 'child neglect'/exp OR abuse:ti,ab OR abused:ti,ab OR abusive:ti,ab OR abusing:ti,ab OR abuser:ti,ab OR abusers:ti,ab OR maltreatment:ti,ab OR maltreated:ti,ab OR neglect:ti,ab OR neglected:ti,ab OR neglecting:ti,ab OR neglects:ti,ab OR "non accidental injury":ti,ab OR "non accidental injuries":ti,ab OR "non accidental trauma":ti,ab OR "non accidental traumas":ti,ab OR "nonaccidental injury":ti,ab OR "nonaccidental injuries":ti,ab OR "nonaccidental trauma":ti,ab OR "nonaccidental traumas":ti,ab OR "non-accidental injury":ti,ab OR "non-accidental injuries":ti,ab OR "non-accidental trauma":ti,ab OR "non-accidental traumas":ti,ab | 277141 |
| 4 | 1 AND 2 AND 3 | 576 |
| 5 | AND [humans]/lim | 565 |

Database (including vendor/platform): Scopus
| Set # |  | Results |
| --- | --- | --- |
| 1 ICD | TITLE-ABS-KEY("International Classification of Diseases" OR ((ICD OR "International Classification of Diseases") AND (codes OR code OR coding OR coded)))) | 32429 |
| 2 Peds | TITLE-ABS-KEY(Infant OR infants OR infancy OR newborn OR newborns OR neonatal OR neonate OR neonates OR baby OR babies OR preterm OR prematurity OR toddler OR toddlers OR boy OR boys OR boyhood OR girl OR girls OR girlhood OR kid OR kids OR child OR childhood OR children OR schoolchild OR schoolgirl OR schoolgirls OR schoolboy OR schoolboys OR "school age" OR "school aged" OR preadolescent OR preadolescents OR preadolescence OR adolescent OR adolescents OR adolescence OR juvenile OR juveniles OR youth OR youths OR teen OR teens OR teenager OR teenagers OR teenaged OR puberty OR pubescent OR pubescence OR prepubescent OR prepubescence OR pediatric OR pediatrics OR paediatric OR paediatrics OR minors) | 6205170 |
| 3 abuse | TITLE-ABS-KEY(abuse OR abused OR abusive OR abusing OR abuser OR abusers OR maltreatment OR maltreated OR neglect OR neglected OR neglecting OR neglects OR "non accidental injury" OR "non accidental injuries" OR "non accidental trauma" OR "non accidental traumas" OR "nonaccidental injury" OR "nonaccidental injuries" OR "nonaccidental trauma" OR "nonaccidental traumas" OR "non-accidental injury" OR "non-accidental injuries" OR "non-accidental trauma" OR "non-accidental traumas") | 581887 |
| 4 | 1 AND 2 AND 3 | 699 |

Database (including vendor/platform): CINAHL
| Set # |  | Results |
| --- | --- | --- |
| 1 ICD | (MH "International Classification of Diseases") OR ((TI ICD OR AB ICD OR TI "International Classification of Diseases" OR AB "International Classification of Diseases") AND (TI codes OR AB codes OR TI code OR AB code OR TI coding OR AB coding OR TI coded OR AB coded)) | 16,463 |
| 2 Peds | MH "Adolescence+" OR MH "Child+" OR MH "Pediatrics+" OR MH "Minors (Legal)" OR MH "Puberty+" OR TI(Infant OR infants OR infancy OR newborn OR newborns OR neonatal OR neonate OR neonates OR baby OR babies OR preterm OR prematurity OR toddler OR toddlers OR boy OR boys OR boyhood OR girl OR girls OR girlhood OR kid OR kids OR child OR childhood OR children OR schoolchild OR schoolgirl OR schoolgirls OR schoolboy OR schoolboys OR "school age" OR "school aged" OR preadolescent OR preadolescents OR preadolescence OR adolescent OR adolescents OR adolescence OR juvenile OR juveniles OR youth OR youths OR teen OR teens OR teenager OR teenagers OR teenaged OR "young person" OR "young persons" OR "young people" OR puberty OR pubescent OR pubescence OR prepubescent OR prepubescence OR pediatric OR pediatrics OR paediatric OR paediatrics OR minors) OR AB(Infant OR infants OR infancy OR newborn OR newborns OR neonatal OR neonate OR neonates OR baby OR babies OR preterm OR prematurity OR toddler OR toddlers OR boy OR boys OR boyhood OR girl OR girls OR girlhood OR kid OR kids OR child OR childhood OR children OR schoolchild OR schoolgirl OR schoolgirls OR schoolboy OR schoolboys OR "school age" OR "school aged" OR preadolescent OR preadolescents OR preadolescence OR adolescent OR adolescents OR adolescence OR juvenile OR juveniles OR youth OR youths OR teen OR teens OR teenager OR teenagers OR teenaged OR "young person" OR "young persons" OR "young people" OR puberty OR pubescent OR pubescence OR prepubescent OR prepubescence OR pediatric OR pediatrics OR paediatric OR paediatrics OR minors) | 1,309,558 |
| 3 abuse | (MH "Child Abuse+") OR TI abuse OR AB abuse OR TI abused OR AB abused OR TI abusive OR AB abusive OR TI abusing OR AB abusing OR TI abuser OR AB abuser OR TI abusers OR AB abusers OR TI maltreatment OR AB maltreatment OR TI maltreated OR AB maltreated OR TI neglect OR AB neglect OR TI neglected OR AB neglected OR TI neglecting OR AB neglecting OR TI neglects OR AB neglects OR (TI "non accidental injury" OR AB "non accidental injury") OR (TI "non accidental | 87611 |
|  | injuries" OR AB "non accidental injuries") OR (TI "non accidental trauma" OR AB "non accidental trauma") OR (TI "non accidental traumas" OR AB "non accidental traumas") OR (TI "nonaccidental injury" OR AB "nonaccidental injury") OR (TI "nonaccidental injuries" OR AB "nonaccidental injuries") OR (TI "nonaccidental trauma" OR AB "nonaccidental trauma") OR (TI "nonaccidental traumas" OR AB "nonaccidental traumas") OR (TI "non-accidental injury" OR AB "non-accidental injury") OR (TI "non-accidental injuries" OR AB "non-accidental injuries") OR (TI "non-accidental trauma" OR AB "non-accidental trauma") OR (TI "non-accidental traumas" OR AB "non-accidental traumas") |  |
| 4 | 1 AND 2 AND 3 | 175 |

Database (including vendor/platform): PsycINFO
| Set # |  | Results |
| --- | --- | --- |
| 1 ICD | DE "International Classification of Diseases" OR ((TI ICD OR AB ICD OR TI "International Classification of Diseases" OR AB "International Classification of Diseases") AND (TI codes OR AB codes OR TI code OR AB code OR TI coding OR AB coding OR TI coded OR AB coded)) | 3804 |
| 2 Peds | DE "Pediatrics" OR TI(Infant OR infants OR infancy OR newborn OR newborns OR neonatal OR neonate OR neonates OR baby OR babies OR preterm OR prematurity OR toddler OR toddlers OR boy OR boys OR boyhood OR girl OR girls OR girlhood OR kid OR kids OR child OR childhood OR children OR schoolchild OR schoolgirl OR schoolgirls OR schoolboy OR schoolboys OR "school age" OR "school aged" OR preadolescent OR preadolescents OR preadolescence OR adolescent OR adolescents OR adolescence OR juvenile OR juveniles OR youth OR youths OR teen OR teens OR teenager OR teenagers OR teenaged OR "young person" OR "young persons" OR "young people" OR puberty OR pubescent OR pubescence OR prepubescent OR prepubescence OR pediatric OR pediatrics OR paediatric OR paediatrics OR minors) OR AB(Infant OR infants OR infancy OR newborn OR newborns OR neonatal OR neonate OR neonates OR baby OR babies OR preterm OR prematurity OR toddler OR toddlers OR boy OR boys OR boyhood OR girl OR girls OR girlhood OR kid OR kids OR child OR childhood OR children OR schoolchild OR schoolgirl OR schoolgirls OR schoolboy OR schoolboys OR "school age" OR "school aged" OR preadolescent OR preadolescents OR preadolescence OR adolescent OR adolescents OR adolescence OR juvenile OR juveniles OR youth OR youths OR teen OR teens OR teenager OR teenagers OR teenaged OR "young person" OR "young persons" OR "young people" OR puberty OR pubescent OR pubescence OR prepubescent OR prepubescence OR pediatric OR pediatrics OR paediatric OR paediatrics OR minors | 1,370,466 |
| 3 abuse | DE "Child Abuse" OR DE "Battered Child Syndrome" OR DE "Child Neglect" OR TI abuse OR AB abuse OR TI abused OR AB abused OR TI abusive OR AB abusive OR TI abusing OR AB abusing OR TI abuser OR AB abuser OR TI abusers OR AB abusers OR TI maltreatment OR AB maltreatment OR TI maltreated OR AB maltreated OR TI neglect OR AB neglect OR TI neglected OR AB neglected OR TI neglecting OR AB neglecting OR TI neglects OR AB neglects OR (TI "non accidental injury" OR AB "non accidental injury") OR (TI "non accidental injuries" OR AB | 174,959 |
|  | "non accidental injuries") OR (TI "non accidental trauma" OR AB "non accidental trauma") OR (TI "non accidental traumas" OR AB "non accidental traumas") OR (TI "nonaccidental injury" OR AB "nonaccidental injury") OR (TI "nonaccidental injuries" OR AB "nonaccidental injuries") OR (TI "nonaccidental trauma" OR AB "nonaccidental trauma") OR (TI "nonaccidental traumas" OR AB "nonaccidental traumas") OR (TI "non-accidental injury" OR AB "non-accidental injury") OR (TI "non-accidental injuries" OR AB "non-accidental injuries") OR (TI "non-accidental trauma" OR AB "non-accidental trauma") OR (TI "non-accidental traumas" OR AB "non-accidental traumas") |  |
| 4 | 1 AND 2 AND 3 | 137 |

